# Echocardiographic and Laboratory Findings in Coronary Slow Flow Phenomenon: cross-sectional study and review

**DOI:** 10.1101/2020.07.03.20145995

**Authors:** Mir Hosein Seyyed Mohammadzad, Salar Gardeshkhah, Kamal Khademvatani, Amin Sedokani

**Author notes:** **Correspondence:** Amin Sedokani, Medical Doctor, Cardiology Department, Medical Faculty, Urmia University of Medical Sciences, Urmia, IRAN., Address: 17 Sharivar St., Urmia, IRAN (Postal Code: 571478334).

## Abstract

**Background and aims:** Coronary Slow flow is a phenomenon known as slow contrast flow that injected into the coronary arteries, without epicardial coronary arteries obstruction. The etiology of this disease is unknown. Endothelial dysfunction, known as a major cause of Coronary Slow flow syndrome (CSF).

**Methods:** This study was cross-sectional (descriptive-analytic), which was performed on patients admitted to Seyedoshohada Heart Center, during one year (2018-2019). Considering the inclusion and exclusion criteria, patients were divided into two groups of normal coronary arteries (NECA, as the control group) and with the Coronary slow-flow (CSF).

**Results:** In the present study, 124 patients were studied, 67.9% in the coronary slow flow group and 39.4% in the control group were men (p=0.001). In the coronary slow flow group, the mean age of patients was 52.18 ± 12.55 years and in the control group, the mean age was 51.77 ± 10.36 years (p=0.18). Mean BMI was significantly higher in the coronary slow flow group than the control group (p <0.05). The smoking, hypertension, and mean of Lymphocyte, Hb, Hct, Plt, MPV, RDW, BUN, FBS, TG, TC, LDL was significantly higher coronary slow flow group. Also, in echocardiography, the mean E wave, E/A ratio was significantly lower in the coronary slow flow group. The GLS was also significantly lower in the control group (p=0.01). LAD was the most common type of coronaries that involved with a slow flow.

**Conclusion:** The results of this study showed that there was a significant increase in the rate of coronary slow flow in men, smokers, high BMI, and hypertensive patients. Also, platelet count, MPV, LDL, FBS, and some laboratory variables were high in patients with CSF. Mild diastolic dysfunction and low GLS were observed in this group of patients.

## 1. Introduction

The coronary slow flow phenomenon (CSFP or cardiac syndrome Y) refers to delayed distal vessel contrast filling in angiography in normal or near-normal coronary arteries that may present in one or all of coronary arteries (1-3). The etiology of the disease is unknown, but histological studies of myofibers hypertrophy, thickening of small vascular muscles with swelling, endothelial cell degradation, and vascular lumen narrowing in these patients (4, 5). The CSFP is diagnosed by Thrombolysis in Myocardial Infarction (TIMI) or the Corrected TIMI Frame Count (CTFC) (6, 7). With advances in transthoracic echocardiography, the pattern of LAD^a^ artery flow can be determined without invasive measures. The accuracy of this method is more than 92% for LAD flow. Patients with CSFP have low diastolic velocity in LAD. This method can also be used to follow up on the treatment of patients (8-10). The prevalence of CSFP is 1- 7% of angiographies. 4% of patients with unstable angina are CSF^b^ patients. The incidence of CSFP is also higher in young men and smokers (5, 11). These patients suffer from recurrent chest pain, frequent hospitalization, and repeated cardiac catheterization. Most importantly, life-threatening arrhythmias (sudden QTc^c^ dispersion) have been reported in these patients. CSF can also lead to myocardial ischemia and acute coronary syndrome (12-14).

Despite many studies, there is no certain explanation for the exact mechanism of CSFP (1-4, 13, 15). It has been reported that there is a decrease in flow-mediated dilatation (FMD) of brachial artery endothelium in CSF patients indicating endothelial dysfunction (16). Increased plasma homocysteine and dimethylarginine levels have also been observed in these patients, which in association with decreased nitric oxide (NO) levels may impair endothelial function (17, 18). Studies have shown that the metabolic syndrome was higher in CSF patients with high cholesterol, high fasting glucose, and high BMI. Insulin resistance and impaired glucose tolerance have also been reported in these patients (19, 20). Another possible cause of this syndrome is the role of the small vessels that are resistive and have a diameter below 400^μm^. These vessels control myocardial blood flow without clear stenosis in the epicardial artery. Some studies have suggested fibromuscular hyperplasia, medial hypertrophy, myo-intimal proliferation, endothelial edema, etc. as possible causes of microvascular dysfunction seen in patients with CSF (5). The use of IVUS^d^ showed that in CSF patients, there was an increase in intimal thickness and diffuse coronary calcification. These findings suggest that CSFP may be a non-obstructive atherosclerotic disease that needs further investigation (21). Also, inflammation that manifests itself with increased CRP, IL-6, WBC, etc. and anatomical factors (geometrical motions, coronary angles, bifurcation, etc.) can cause CSFP (22, 23).

Two-dimensional Speckle Tracking Echocardiography (STE) is a new imaging method that, like tissue Doppler imaging (TDI), can calculate offline myocardial velocities and deformation parameters such as strain rate (SR). the global longitudinal strain (GLS) is also a robust, valid and repeatable technique for measuring left ventricular deformation by echocardiography. The GLS appears to be a sensitive method for the subclinical investigation of myocardial abnormalities that can analyze and treat regional and generalized wall defects (24, 25). Recent studies have shown that the study of total wall tension (GLS) by the STE method can even reveal the most minor wall changes in people with normal LVEF (26). Altunkas et al. showed that left ventricular systolic and diastolic function decreased in patients with the CSF while no change in right ventricular function was observed and these changes can be monitored by the TDI/GLS method (27). In other studies, with emphasis on TDI/GLS evaluation in patients with the CSF, they reported impaired left ventricular systolic and diastolic function in patients with CSF (by 2D/TDI) (28). Various studies have examined some aspects of the involvement in CSF, for example, studies of the TDI/GLS echocardiography and some other studies of clinical and laboratory predictors, risk factors, angiographic profile, and clinical findings in CSFP (29-32). But so far, no study has comprehensively investigated the laboratory and echocardiography findings of CSF phenomenon in angiography.

In the present study, we aimed to investigate the comprehensive echocardiographic, and laboratory findings in the patients with coronary slow flow phenomenon in angiography.

## 2. Material and Methods

This study is a cross-sectional (descriptive-analytical) study that performed in a year (August 2018- August 2019) in Seyedoshohada heart center of West Azarbayjan province of Iran and 124 patients enrolled in the study. Patient information was recorded in a pre-prepared checklist. Taking into account the conditions of entry and exit of patients was divided into two separate groups of normal epicardial coronary artery (NECA**-**as the control group) and coronary slow flow (CSF) and then in the same conditions for both groups, the necessary information for each group was separately completed.

In the present study, patients with normal systolic function entered the study. The tests have been registered based on variables (such as WBC, Hb, Plt, MPV, RDW, Cr, FBS, TG, TC, LDL, and HDL) in a pre-prepared checklist. Patients were also subjected to echocardiography and their information was recorded separately in a standard checklist previously prepared (including demographic, age, sex, paraclinical and clinical information on underlying diseases, type of vessel involved, etc.). The instrument for diagnosing and evaluating SCF patients was coronary angiography, with TIMI greater than 27 and TIMI-2-Flow, which are quantitative and qualitative methods of assessing coronary blood flow, respectively (6, 11). Besides, after providing complete explanations about the study to the participants, they were given informed consent and all information remained confidential.

Exclusion criteria for the patients were the previous history of coronary heart disease (including the history of stent implantation, history of CABG surgery, and atherosclerotic coronary stenosis on a previous angiography), any significant valvular heart disease, pulmonary hypertension, history of chronic obstructive pulmonary disease (COPD), and atrioventricular conduction disturbances.

### 2.1. Data Analysis

The obtained information was expressed as mean, frequency and percentage. For qualitative variables such as smoking, sex, family history, percentage, etc., and for quantitative variables such as height, BMI, laboratory variables, mean±SD were used. The quantitative variables were compared using Student t-test (Independent Samples) or Mann-Whitney U test (depending on the type of data distribution). The qualitative variables were analyzed using Chi-Square Test by SPSS V.26 software. In all of the test, the significance level was P <0.05.

### 2.2. Ethical statement

All of the stages of the study were under supervision and confirmation of the ethical committee of Seyedoshohada heart center and Urmia University of Medical Sciences. After all the patients were provided with written and oral explanations, they received written consent (informed written consent was given prior to the inclusion of subjects in the study). All patients had the opportunity to withdraw from the study at any time. Patient information remained confidential. There was not any intervention or extra cost to patients. The special approved ethical code for the study from ethics committee of Ministry of Health of Iran is <u>IR.UMSU.REC.1397.236</u>. The investigation has met the principles outlined in the Declaration of Helsinki.

## 3. Results

### 4.1. Gender

In the present study, 124 patients were examined that in the CSF group, 67.9% (36 people) were male and 32.1% (17 people) were female, and in the control group, 39.4% (26 people) were male and 60.6% (43 people) are women. Test results showed that there was a significant relationship between coronary artery blood flow and sex, and the frequency of men in the coronary artery blood flow group was higher than the control group (p=0.001).

### 4.2. Age

In the CSF group, the average age of the patients 55.12±18.52 years, the youngest patient was 26 and the oldest one is 74 years old. In the control group, the mean age was 51.77±10.36 years, the youngest person was 27 years old and the oldest was 72 years old. There was no significant difference between the mean age in the coronary blood flow group and the control group (p=0.18).

### 4.3. Family History of Coronary Heart Disease

Our results indicated that, in the CSF group, 35.8% and in NECA, 21.1% had a positive family history for coronary heart disease, but the difference was not significant (p=0.10).

### 4.4. Body Mass Index

In the CSF group, mean BMI equal to 28.13±2.28 kg/m^2^, the lowest amount equal to 23 and the maximum amount equal to 33 kilograms per square meter. In the control group, the mean BMI was 24.58±1.64 kg/m^2^, the lowest value was 21 and the highest value was 29 kg/m^2^. The mean BMI in the CSF group was significantly higher than the control group (P <0.001).

### 4.5. Smoking

In the CSF group, 62.2% (33 patients) of patients were active smokers while in the NECA group this ratio was 18.3% (13 patients). The active smokers were more frequent in the CSF group significantly (P <0.001).

### 4.6. Hypertension

The frequency of HTN among CSF and control group patients were as 58.5% and 25.4% respectively. The results indicated that HTN in CSF patients is significantly more than NECA patients (P <0.001).

### 4.7. Diabetes, Dyslipidemia, and Chronic Kidney Diseases

In the assessment of diabetes mellitus, dyslipidemia, and CKD, our results indicated that there were no significant differences in the mentioned diseases with CSF phenomenon among groups for the p-values were 0.91, 0.49, and 0.65 respectively.

### 4.8. Laboratory Findings

An independent t-test was used to investigate the association of coronary artery flow with laboratory parameters. The results show that the mean of Neutrophil absolute count, Lymphocyte absolute count, Hct, Plt, MPV, RDW, Cr, TG, TC, LDL in the CSF blood flow group is significantly higher than the control group (Table 1, Figure 1).

**Figure 1.**
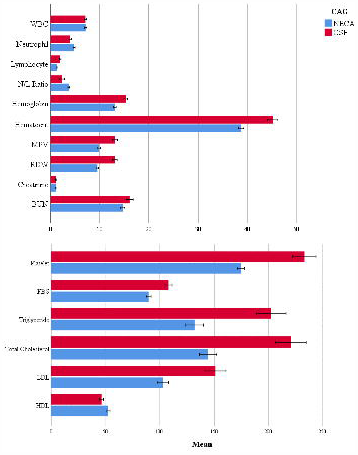
Laboratory data of the Coronary Slow Flow (53 patients) and Normal Epicardial Coronary Artery Groups (71 patients). Student t-test, Error Bar Confidence Interval: 95%.

**Figure 2.**
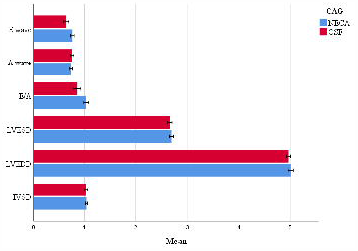
Echocardiographic findings of the Coronary Slow Flow (53 patients) and Normal Epicardial Coronary Artery Groups (71 patients). Student t-test, Error Bar Confidence Interval: 95%.

**Figure 3.**
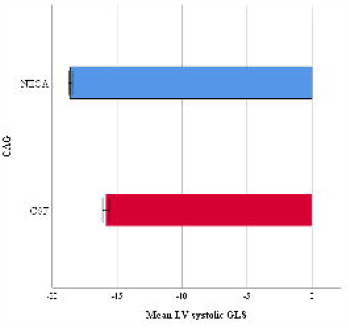
Mean Left Ventricle Global Longitudinal Strain Comparison of the Coronary Slow Flow (53 patients) and Normal Epicardial Coronary Artery Groups (71 patients). Student t-test, Error Bar Confidence Interval: 95%.

### 4.9. Echocardiographic Findings

An independent t-test was also used to investigate the association between coronary slow blood flow and echocardiographic parameters. Our results showed that LVEF, IVSD, E wave, and LV systolic GLS are significantly higher in the NECA group (Table 1).

### 4.10. Artery Type

In our study, 46.8% of the CSF arteries were on LAD, 27.9% were on LCX and 25.3% were on RCA. Due to this LAD was the most common artery with CSF phenomenon.

### 4.11. Global Longitudinal Strain for LV Function

The results of the independent t-test show that the mean LV systolic GLS is significantly higher in the NECA group than the CSF group (p=0.01, Table 1).

## 4. Discussion

The CSF blood flow is an angiographic diagnosis of slow-flowing contrast material in normal or near-normal vessels in coronary epicardial vessels (1-3). The CSF mimics a variety of clinical manifestations, including unstable angina, acute myocardial infarction, and ventricular tachycardia (33). The exact cause and mechanism of its pathophysiology are not yet known, but various studies have suggested various hypotheses, including endothelial dysfunction, microvascular dysfunction, early-stage coronary atherosclerosis, metabolic disorders, and inflammatory disease (34-39). Researchers have observed in biopsies of patients with CSF, fibromuscular hyperplasia, medial hypertrophy, myointimal proliferation, endothelial edema, and degeneration in small vessels (5, 34). At present, obstructive coronary artery disease has been suggested as a possible cause (5). It has recently been reported that decreased adiponectin concentrations and the activity of paraoxonase, two important markers of endothelial dysfunction, are associated with CSF (40).

In the present study, in the CSF group, the mean age of the patients was higher than the control group. Also, in the CSF group, most of the patients were male, and in terms of gender, the prevalence of men was significantly higher in this group than in the control group. In many similar studies, no significant difference in age was observed among patients, but in terms of prevalence, the CSF was higher in men (28-32, 41, 42). It can be said that based on the present study and other studies, age cannot affect the mechanism of the CSF phenomenon, or at least more detailed information can be provided by conducting further studies with a long research period and several angiographic studies. Gender-oriented pathophysiology has not yet been proposed. It may be due to hormonal changes, stressful situations, or different job situations.

In this study, BMI was significantly higher in patients with the CSF than in the control group, and a decrease in BMI could reduce the incidence of CSF in patients. The results obtained are almost similar to the results of most studies conducted in this field (29, 30, 32, 42). Although Sanati et al.’s study reported that low BMI was associated with the CSFP (42). But other studies, including Yilmaz et al., in a study of Turkish society, showed that high BMI had an independent association with the CSF (19), and similarly Hawkins et al. showed that BMI was an independent predictor for the CSF in the North American population (43). Obesity can lead to endothelial dysfunction and can play a role in the CSF pathogenesis, as Pontiroli et al, showed that after gastric banding procedure, BMI decreased and improved endothelial system function (44). BMI is a predictor of CSF in the population, so other studies are needed to show a link between lowering BMI and improving CSF.

In the present study, smoking and high blood pressure were significantly higher in patients with the CSF, but no difference was observed between the two groups in terms of diabetes, dyslipidemia, and family history of coronary heart disease. In some studies, there was no significant difference between the two groups in terms of diabetes, blood pressure, dyslipidemia and family history of coronary heart disease (32), while in others there was a significant relationship in terms of smoking, blood pressure or the frequency of smokers and people with high blood pressure in patients with CSF (28, 29, 41, 42)

The risk factors or pathophysiology of the CSF have not yet been fully elucidated, and indifferent studies based on different populations and cultures; they are exposed to various risk factors. As the results of the above studies confirmed. In one study in Australia, the most important risk factors for the CSF were smoking and male sex (2). However, in another study in China, hyperuricemia, hyperglycemia, and high hsCRP have been identified as factors that cause endothelial dysfunction as independent risk factors for the CSF (45). In the study of Moazenzadeh et al, diabetes, hypertension, and opioid use were risk factors for the CSF (46). Different populations (genetic differences) have different risk factors for the occurrence of the CSF, so it can be said that one or more of these risk factors may play a role in the development of the CSF by causing endothelial system disorders.

The results of the present study showed that the mean of Lymphocyte, Hb, Hct, Plt, MPV, RDW, BUN, FBS, TG, Total Cholesterol, LDL in the CSF blood flow group was significantly higher than the control group. While the mean Neutrophil and HDL in the coronary artery blood flow group were significantly lower than the control group and there was no significant difference in terms of WBC and Creatinine. Laboratory results, such as risk factors, can vary from patient to patient.

In Yildiz et al.’s study, creatinine, uric acid, hematocrit, MCV, and HDL levels were lower, but there was no significant difference between WBC, platelets, fasting blood sugar, and MPV (28, 33, 42). In the study by Ghaffari et al., total cholesterol, triglycerides, hemoglobin, hematocrit, lymphocytes count, platelet count, platelet distribution, RDW, MPV, and fasting blood sugar were also significantly higher in patients with the CSF. were higher in the CSF group, but there was no difference in WBC, creatinine, monocytes, and neutrophils between groups. Finally, it has been reported that people with the CSF may have an underlying inflammation and endothelial dysfunction (29). Previous studies and studies by Narimani et al. have highlighted high LDL and low HDL levels in patients with the CSF blood flow (32), indicating that by modifying the fat profile through diet, physical activity, or medication, the incidence of the CSF can be reduced. However, like most of its mechanism is not clear at present. In some other studies, there was no significant difference in hematocrit, platelet, uric acid, HbA1C, homocysteine (30), and in general lipid profile and blood markers (31). However, we need more research in this area.

In a study by Altun et al., the results showed that creatinine and hemoglobin levels were high in CSF patients. MPV, RDW, and neutrophil-to-lymphocyte (N/L) ratios did not differ significantly between the two groups. Inflammation has been shown to play an important role in the CSF pathogenesis. However, the association of the CSF with inflammatory markers is controversial (41). In some other studies, there has been a strong association between RDW and inflammatory markers (47). Studies have also shown that high RDW is associated with the CSF (48).

Platelet dysfunction can also play a role in the CSF pathogens. Gokce et al. Showed that platelet aggregation was higher in patients with the CSF than in controls. Therefore, it can be suggested that one of the possible causes of the pathophysiology of the CSF can be platelet dysfunction. The size of the platelets shown by the MPV is a useful marker for platelet function. And it has been shown that MPV levels also have a significant positive association with the CSF (49, 50).

Examining the association of the CSF blood flow with echocardiographic parameters, the results of the present study show that the mean, E wave, and E/A ratio in the coronary blood flow group are significantly lower and DT is higher than the control group, which can indicate mild diastolic disorder in patients with the CSF. In Wang et al.’s study, the E/A ratios and E waves were significantly reduced in CSF patients, yet there was no significant difference in terms of wave A (28). Also, Baykan et al reported left ventricular systolic and diastolic disturbances in patients with the CSF (51). In some studies, it has been suggested that in patients with the CSFP, ischemic coronary artery disease may develop systolic and diastolic left ventricular dysfunction (52-54). However, in the present study, the only diastolic disturbance was observed and there was no significant difference in terms of EF. However, the GLS of patients with the CSF was lower, which could indicate physiological left ventricular dysfunction, which will be discussed below. The levels of LVEF, IVSD, LVESD, LVEDD, and A wave did not differ significantly between the CSF and the NECA groups. In the study of Narimani et al. In terms of LVESD, LVEDD, EF, E and A waves, E/A ratio, DT, IVRT, there was no significant difference between the two groups, but e and s’ lateral wall waves were lower in patients the CSF (32) but in another study, there was no difference in LVEF (28).

By measuring GLS, we are able to measure myocardial movements in all directions, and this criterion gives us the amount of left ventricular tension at all angles (longitudinal, radial, and marginal). This completely non-invasive method is very helpful in diagnosing and evaluating systolic and diastolic myocardial function (55). GLS is a sensitive device for diagnosing left ventricular dysfunction at the onset of many myocardial pathological conditions (39, 56). In the present study, the mean GLS in the CSF blood flow group was significantly lower than the control group. Wang et al. Reported that left ventricular diastolic and systolic longitudinal traction was lower in patients with the CSF than in the control group (28). However, in the study of Narimani et al., No statistically significant relationship was observed in terms of systolic and diastolic longitudinal traction and showed that the CSF could not impair systolic and diastolic longitudinal function (32). Nurkalem and colleagues also showed that the left ventricular and left ventricular longitudinal strain was different between the two groups (57). There are controversies in left ventricular function in patients with CSF, so further studies are needed. It has previously been shown that left ventricular longitudinal traction impairment is affected earlier than marginal and radial traction (58).

In the present study, LAD was more common in terms of abundance of coronary arteries in terms of the CSF, followed by LCX and RCA. In Yildiz et al.’s study, the prevalence of the CSF in the LAD artery was also higher (42). Also, in other similar studies, LAD, LCX, and RCA had the highest prevalence in terms of the prevalence of the CSF (28, 30, 31). The prevalence of the CSF in different arteries can vary widely in different technical and racial contexts, and the variable involved in each vascular study. However, LAD was the most prevalent in most studies.

The results of the present study showed that in men, smokers, patients with high BMI, and hypertension, the prevalence of Coronary Slow Flow was significantly high. Also, platelet counts, MPV, LDL, FBS and some laboratory variables were high in patients with slow coronary flow. In addition, in echocardiography, mild diastolic disturbance and decreased left ventricular GLS were found in the Coronary Slow Flow patients.

## Data Availability

Not available for public

## 6. All of authors report no kind of conflict of interests in this study and there is nothing

## 8. Author contributions

All of the authors in this study have contributed equally in design, performance, data collection, analysis, and writing and review of the manuscript.

## 9. Acknowledgements

We would like to thank the health care personnel of Seyedoshohada heart center, specially the angiography and echocardiography ward personnel.

## Footnotes

a Left Anterior Descending artery

b Coronary Slow Flow

c Corrected QT Interval

d Intravascular Ultrasonography

## References

1. Kopetz V, Kennedy J, Heresztyn T, Stafford I, Willoughby SR, Beltrame JF. Endothelial function, oxidative stress and inflammatory studies in chronic coronary slow flow phenomenon patients. Cardiology. 2012;121(3):197–203.

2. Beltrame JF, Limaye SB, Horowitz JD. The coronary slow flow phenomenon--a new coronary microvascular disorder. Cardiology. 2002;97(4):197–202.

3. Tambe AA, Demany MA, Zimmerman HA, Mascarenhas E. Angina pectoris and slow flow velocity of dye in coronary arteries--a new angiographic finding. Am Heart J. 1972;84(1):66–71.

4. Caglayan AO, Kalay N, Saatci C, Yalcyn A, Akalyn H, Dundar M. Lack of association between the Glu298Asp polymorphism of endothelial nitric oxide synthase and slow coronary flow in the Turkish population. Can J Cardiol. 2009;25(3):e69–72.

5. Mosseri M, Yarom R, Gotsman MS, Hasin Y. Histologic evidence for small-vessel coronary artery disease in patients with angina pectoris and patent large coronary arteries. Circulation. 1986;74(5):964–72.

6. Gibson CM, Cannon CP, Daley WL, Dodge JT, Jr., Alexander B, Jr., Marble SJ, et al. TIMI frame count: a quantitative method of assessing coronary artery flow. Circulation. 1996;93(5):879–88.

7. Kaski JC, Eslick GD, Merz CNB. Chest Pain with Normal Coronary Arteries: A Multidisciplinary Approach: Springer Science & Business Media; 2013.

8. Citro R, Voci P, Pizzuto F, Maione AG, Patella MM, Bossone E, et al. Clinical value of echocardiographic assessment of coronary flow reserve after left anterior descending coronary artery stenting in an unselected population. J Cardiovasc Med (Hagerstown). 2008;9(12):1254–9.

9. Tani T, Tanabe K, Kitai T, Yamane T, Kureha F, Katayama M, et al. Detection of severe stenosis and total occlusion in the left anterior descending coronary artery with transthoracic Doppler echocardiography in the emergency room. Echocardiography. 2009;26(1):15–20.

10. Anjaneyulu A, Raghu K, Chandramukhi S, Satyajit GM, Arramraja S, Raghavaraju P, et al. Evaluation of left main coronary artery stenosis by transthoracic echocardiography. J Am Soc Echocardiogr. 2008;21(7):855–60.

11. Diver DJ, Bier JD, Ferreira PE, Sharaf BL, McCabe C, Thompson B, et al. Clinical and arteriographic characterization of patients with unstable angina without critical coronary arterial narrowing (from the TIMI-IIIA Trial). Am J Cardiol. 1994;74(6):531–7.

12. Beltrame JF, Limaye SB, Wuttke RD, Horowitz JD. Coronary hemodynamic and metabolic studies of the coronary slow flow phenomenon. Am Heart J. 2003;146(1):84–90.

13. Li JJ, Xu B, Li ZC, Qian J, Wei BQ. Is slow coronary flow associated with inflammation? Med Hypotheses. 2006;66(3):504–8.

14. Cutri N, Zeitz C, Kucia AM, Beltrame JF. ST/T wave changes during acute coronary syndrome presentation in patients with the coronary slow flow phenomenon. Int J Cardiol. 2011;146(3):457–8.

15. Sezgin AT, Sigirci A, Barutcu I, Topal E, Sezgin N, Ozdemir R, et al. Vascular endothelial function in patients with slow coronary flow. Coron Artery Dis. 2003;14(2):155–61.

16. Spiro JR, Digby JE, Ghimire G, Mason M, Mitchell AG, Ilsley C, et al. Brachial artery low-flow-mediated constriction is increased early after coronary intervention and reduces during recovery after acute coronary syndrome: characterization of a recently described index of vascular function. Eur Heart J. 2011;32(7):856–66.

17. Riza Erbay A, Turhan H, Yasar AS, Ayaz S, Sahin O, Senen K, et al. Elevated level of plasma homocysteine in patients with slow coronary flow. Int J Cardiol. 2005;102(3):419–23.

18. Selcuk MT, Selcuk H, Temizhan A, Maden O, Ulupinar H, Baysal E, et al. Asymmetric dimethylarginine plasma concentrations and L-arginine/asymmetric dimethylarginine ratio in patients with slow coronary flow. Coron Artery Dis. 2007;18(7):545–51.

19. Yilmaz H, Demir I, Uyar Z. Clinical and coronary angiographic characteristics of patients with coronary slow flow. Acta Cardiol. 2008;63(5):579–84.

20. Ozcan T, Gen R, Akbay E, Horoz M, Akcay B, Genctoy G, et al. The correlation of thrombolysis in myocardial infarction frame count with insulin resistance in patients with slow coronary flow. Coron Artery Dis. 2008;19(8):591–5.

21. Cin VG, Pekdemir H, Camsar A, Cicek D, Akkus MN, Parmaksyz T, et al. Diffuse intimal thickening of coronary arteries in slow coronary flow. Jpn Heart J. 2003;44(6):907–19.

22. Kalay N, Aytekin M, Kaya MG, Ozbek K, Karayakali M, Sogut E, et al. The relationship between inflammation and slow coronary flow: increased red cell distribution width and serum uric acid levels. Turk Kardiyol Dern Ars. 2011;39(6):463–8.

23. Kantarci M, Gundogdu F, Doganay S, Duran C, Kalkan ME, Sagsoz ME, et al. Arterial bending angle and wall morphology correlate with slow coronary flow: determination with multidetector CT coronary angiography. Eur J Radiol. 2011;77(1):111–7.

24. Kalam K, Otahal P, Marwick TH. Prognostic implications of global LV dysfunction: a systematic review and meta-analysis of global longitudinal strain and ejection fraction. Heart. 2014;100(21):1673–80.

25. Kurt M, Tanboga IH, Aksakal E. Two-Dimensional Strain Imaging: Basic principles and Technical Consideration. Eurasian J Med. 2014;46(2):126–30.

26. Kaneko A, Tanaka H, Onishi T, Ryo K, Matsumoto K, Okita Y, et al. Subendocardial dysfunction in patients with chronic severe aortic regurgitation and preserved ejection fraction detected with speckle-tracking strain imaging and transmural myocardial strain profile. Eur Heart J Cardiovasc Imaging. 2013;14(4):339–46.

27. Altunkas F, Koc F, Ceyhan K, Celik A, Kadi H, Karayakali M, et al. The effect of slow coronary flow on right and left ventricular performance. Med Princ Pract. 2014;23(1):34–9.

28. Wang Y, Ma C, Zhang Y, Guan Z, Liu S, Li Y, et al. Assessment of left and right ventricular diastolic and systolic functions using two-dimensional speckle-tracking echocardiography in patients with coronary slow-flow phenomenon. PLoS One. 2015;10(2):e0117979.

29. Ghaffari S, Tajlil A, Aslanabadi N, Separham A, Sohrabi B, Saeidi G, et al. Clinical and laboratory predictors of coronary slow flow in coronary angiography. Perfusion. 2017;32(1):13–9.

30. Mukhopadhyay S, Kumar M, Yusuf J, Gupta VK, Tyagi S. Risk factors and angiographic profile of coronary slow flow (CSF) phenomenon in North Indian population: An observational study. Indian Heart J. 2018;70(3):405–9.

31. Sanati H, Kiani R, Shakerian F, Firouzi A, Zahedmehr A, Peighambari M, et al. Coronary Slow Flow Phenomenon Clinical Findings and Predictors. Res Cardiovasc Med. 2016;5(1):e30296.

32. Narimani S, Hosseinsabet A, Pourhosseini H. Effect of Coronary Slow Flow on the Longitudinal Left Ventricular Function Assessed by 2-Dimensional Speckle-Tracking Echocardiography. J Ultrasound Med. 2016;35(4):723–9.

33. Saya S, Hennebry TA, Lozano P, Lazzara R, Schechter E. Coronary slow flow phenomenon and risk for sudden cardiac death due to ventricular arrhythmias: a case report and review of literature. Clin Cardiol. 2008;31(8):352–5.

34. Mangieri E, Macchiarelli G, Ciavolella M, Barilla F, Avella A, Martinotti A, et al. Slow coronary flow: clinical and histopathological features in patients with otherwise normal epicardial coronary arteries. Cathet Cardiovasc Diagn. 1996;37(4):375–81.

35. Serne EH, de Jongh RT, Eringa EC, Rg IJ, Stehouwer CD. Microvascular dysfunction: a potential pathophysiological role in the metabolic syndrome. Hypertension. 2007;50(1):204–11.

36. Celebi H, Catakoglu AB, Kurtoglu H, Sener M, Hanavdelogullari R, Demiroglu C, et al. The relation between coronary flow rate, plasma endothelin-1 concentrations, and clinical characteristics in patients with normal coronary arteries. Cardiovasc Revasc Med. 2008;9(3):144–8.

37. Pekdemir H, Cin VG, Cicek D, Camsari A, Akkus N, Doven O, et al. Slow coronary flow may be a sign of diffuse atherosclerosis. Contribution of FFR and IVUS. Acta Cardiol. 2004;59(2):127–33.

38. Li JJ, Qin XW, Li ZC, Zeng HS, Gao Z, Xu B, et al. Increased plasma C-reactive protein and interleukin-6 concentrations in patients with slow coronary flow. Clin Chim Acta. 2007;385(1-2):43–7.

39. Mor-Avi V, Lang RM, Badano LP, Belohlavek M, Cardim NM, Derumeaux G, et al. Current and evolving echocardiographic techniques for the quantitative evaluation of cardiac mechanics: ASE/EAE consensus statement on methodology and indications endorsed by the Japanese Society of Echocardiography. J Am Soc Echocardiogr. 2011;24(3):277–313.

40. Selcuk H, Selcuk MT, Temizhan A, Maden O, Saydam GS, Ulupinar H, et al. Decreased plasma concentrations of adiponectin in patients with slow coronary flow. Heart Vessels. 2009;24(1):1–7.

41. Altun I, Akin F, Kose N, Sahin C, Kirli I. Predictors of slow flow in angiographically normal coronary arteries. Int J Clin Exp Med. 2015;8(8):13762–8.

42. Yildiz A, Sezen Y, Gunebakmaz O, Kaya Z, Altiparmak IH, Erkus E, et al. Association of Meteorological Variables and Coronary Blood Flow. Clin Appl Thromb Hemost. 2015;21(6):570–8.

43. Hawkins BM, Stavrakis S, Rousan TA, Abu-Fadel M, Schechter E. Coronary slow flow--prevalence and clinical correlations. Circ J. 2012;76(4):936–42.

44. Pontiroli AE, Pizzocri P, Paroni R, Folli F. Sympathetic overactivity, endothelial dysfunction, inflammation, and metabolic abnormalities cluster in grade III (World Health Organization) obesity: reversal through sustained weight loss obtained with laparoscopic adjustable gastric banding. Diabetes Care. 2006;29(12):2735–8.

45. Xia S, Deng SB, Wang Y, Xiao J, Du JL, Zhang Y, et al. Clinical analysis of the risk factors of slow coronary flow. Heart Vessels. 2011;26(5):480–6.

46. Moazenzadeh M, Azimzadeh BS, Zare J, Shokouhi M, Sheikhvatan M. Clinical features and main determinants of coronary slow flow phenomenon in Iranian patients. Eur J Cardiovasc Med. 2010;1(2):2042.

47. Patel KV, Ferrucci L, Ershler WB, Longo DL, Guralnik JM. Red blood cell distribution width and the risk of death in middle-aged and older adults. Arch Intern Med. 2009;169(5):515–23.

48. Luo SH, Jia YJ, Nie SP, Qing P, Guo YL, Liu J, et al. Increased red cell distribution width in patients with slow coronary flow syndrome. Clinics (Sao Paulo). 2013;68(6):732–7.

49. Gokce M, Kaplan S, Tekelioglu Y, Erdogan T, Kucukosmanoglu M. Platelet function disorder in patients with coronary slow flow. Clin Cardiol. 2005;28(3):145–8.

50. Lanza G, Andreotti F, Sestito A, Sciahbasi A, Crea F, Maseri A. Platelet aggregability in cardiac syndrome X. European heart journal. 2001;22(20):1924–30.

51. Baykan M, Baykan EC, Turan S, Gedikli O, Kaplan S, Kiris A, et al. Assessment of left ventricular function and Tei index by tissue Doppler imaging in patients with slow coronary flow. Echocardiography. 2009;26(10):1167–72.

52. Demirkol MO, Yaymaci B, Mutlu B. Dipyridamole myocardial perfusion single photon emission computed tomography in patients with slow coronary flow. Coron Artery Dis. 2002;13(4):223–9.

53. Buchthal SD, den Hollander JA, Merz CN, Rogers WJ, Pepine CJ, Reichek N, et al. Abnormal myocardial phosphorus-31 nuclear magnetic resonance spectroscopy in women with chest pain but normal coronary angiograms. N Engl J Med. 2000;342(12):829–35.

54. Yaymaci B, Dagdelen S, Bozbuga N, Demirkol O, Say B, Guzelmeric F, et al. The response of the myocardial metabolism to atrial pacing in patients with coronary slow flow. Int J Cardiol. 2001;78(2):151–6.

55. Biering-Sorensen T, Solomon SD. Assessing Contractile Function When Ejection Fraction Is Normal: A Case for Strain Imaging. Circ Cardiovasc Imaging. 2015;8(11):e004181.

56. Yuda S, Fang ZY, Marwick TH. Association of severe coronary stenosis with subclinical left ventricular dysfunction in the absence of infarction. J Am Soc Echocardiogr. 2003;16(11):1163–70.

57. Nurkalem Z, Gorgulu S, Uslu N, Orhan AL, Alper AT, Erer B, et al. Longitudinal left ventricular systolic function is impaired in patients with coronary slow flow. Int J Cardiovasc Imaging. 2009;25(1):25–32.

58. Edvardsen T, Helle-Valle T, Smiseth OA. Systolic dysfunction in heart failure with normal ejection fraction: speckle-tracking echocardiography. Prog Cardiovasc Dis. 2006;49(3):207–14.

